# The first GAEN-based COVID-19 contact tracing app in Norway identifies 80% of close contacts in “real life” scenarios

**DOI:** 10.1101/2021.05.06.21253948

**Authors:** Hinta Meijerink, Elisabeth H. Madslien, Camilla Mauroy, Mia Karoline Johansen, Sindre Møgster Braaten, Christine Ursin Steen Lunde, Trude Margrete Arnesen, Siri Laura Feruglio, Karin Nygård

## Abstract

The COVID-19 response in most countries depends on testing, isolation, contact tracing, and quarantine, which is labor- and time consuming. Therefore, several countries worldwide launched Bluetooth based apps as supplemental tools. We evaluated the new Norwegian GAEN (Google Apple Exposure Notification) based contact tracing app “Smittestopp” under two relevant simulated scenarios, namely standing in a queue and riding public transport.

We compared two configurations (C1: 58/63 dBm; C2: 58/68 dBm) with multiple weights (1.0-2.5) and time thresholds (10-15 min), by calculating notification rates among close contacts (≤2 meters, ≥15 min) and other non-close contacts. In addition, we estimated the effect of using different operating systems and locations of phone (hand/pocket) using χ^2^.

C2 resulted in significantly higher notification rates than C1 (p-value 0.05 - 0.005). The optimal setting resulted in notifications among 80% of close contacts and 34% of other contacts, using C2 with weights of 2.0 for the low and 1.5 for the middle bucket with a 13-minutes time threshold. Among other contacts, the notification rate was 67% among those ≤2 meters for <15 minutes compared to 19% among those >2 meters (p=0.004). Significantly (p-values 0.046 - 0.001) lower notification rates were observed when using the iOS operating systems or carrying the phone in the pocket instead of in the hand.

This study highlights the importance of testing and optimizing the performance of contact tracing apps under “real life” conditions to optimized configuration for identifying close contacts.

## Background

Until March 9th 2020, all cases of coronavirus disease (COVID-19) in Norway were associated with travel or contact with a confirmed case.^1^ When cases with unidentified source of infection were reported, Norway imposed comprehensive control measures, including closure of schools, training facilities, and a variety of businesses, and service industries. The response is based on “Test, Isolate, Trace and Quarantine (TITQ)”; test suspected cases, isolate confirmed cases, identify close contacts (≤2 meters, ≥15 minutes), and quarantine those close contacts.^2^ One key factor of successful TITQ strategy and breaking chains of infection is early identification of contacts. In 2020, Norway reported 50 130 confirmed cases, of which 31 155 were infected in Norway and 4 360 abroad.^3^ Based on the last three months of 2020, when testing capacity allowed testing everyone with symptoms or suspected exposure, in 30% of cases country of infection was unknown and among cases infected in Norway the source of transmission or exposure was unknown or missing in 20%.^3,4^ This indicates that a proportion of cases and contacts are not identified through manual contact tracing or do not follow-up advise given as reported by other countries.^5^

Manual contact tracing is an efficient tool in limiting the spread of COVID-19, but is labor-and time consuming and depends on factors such as, the capacity of the local contact tracing teams, the number of contacts per confirmed case as well as cases knowing their contacts.^6,7^ In Norway, contact tracing has sometimes been hampered by challenges including hesitation to answer anonymous phone calls from the contact tracing teams, lacking or incorrect contact info for contacts (e.g. people with temporary citizenship or tourists), language/cultural barriers, gaps in memory, or general unwillingness to collaborate. Therefore, contact notification might be delayed by days and some contacts might not be identified. Digital solutions, such as Bluetooth based apps, have been proposed as a supplemental tool and have the advantage of providing rapid notification of possible community exposures.^8-10^ This could be particularly helpful in hot-spot areas, where population density is high and social distancing in public spaces is challenging. However, the Bluetooth-based technology still has several limitations in terms of the ability to correctly identify close contacts.^11^ Proximity estimation is based on the decaying/attenuation of Bluetooth signal, measured as attenuation of the Received Signal Strength Indicator (RSSI) values (dBm), which is affected by many factors and not a precise measurement of distance.^11,12^ Furthermore, the success of the app-based approach strongly depends on the population’s acceptance to using it. For the app to be widely accepted, it is essential that people are willing to act upon receiving a notification.^9^ If, for instance, people had to be quarantined for 10 days, without knowing if they fulfilled the criteria for what is defined a “close contact” or not, it is not likely that many would download it or follow the advice.

In April 2020, Norway released an app for COVID-19 contact tracing (Smittestopp v1), which used a centralized approach that registered data from Bluetooth contacts and location into a central database. This solution was intended to fulfill two purposes: notifying close contact of individuals with confirmed COVID-19 infection as well as analyzing movement patterns and population behavior during the pandemic. However, in June 2020, Smittestopp v1 was shut down by the Norwegian Data Protection Authority (Datatilsynet) due to privacy concerns, specifically regarding the centralized storage of positional data and Bluetooth contacts. Subsequently, Norway developed a new app (Smittestopp v2) based on the Google/Apple Exposure Notification (GAEN) API, which uses a decentralized approach and allows international integration.^13,14^

When GAEN is activated on a device, it will generate a random identifier, Temporary Exposure Key (TEK), for each day, and from this generate short lived Rotating Proximity Identifiers (RPI). The RPIs are broadcasted to nearby devices, while the TEK stays on the device. Additionally, the device scans for RPIs from other devices. Consequently, phones exchange RPIs when in proximity, which are stored for 14 days on the phone and can later be used to identify exposure. When someone tests positive for COVID-19, they can report themselves infected via a secure system (ID-porten) which checks if they have been reported in the Norwegian Surveillance System for Communicable Diseases (MSIS). Upon confirmation, the TEKs for the last 14 days are shared as “infected keys” with all other users. These keys are used to identify relevant exposures based on when exposure took place, how long the contact was and the Bluetooth attenuation, based on matching the shared TEKs with RPIs stored on the phone. The Smittestopp v2 was launched Dec 2020 and as of February 1^st^ 2021 was downloaded 691 400 times and 728 infections were reported.^15^

Over 20 countries have implemented GAEN based contact tracing apps, including Germany, Ireland, Denmark, Netherlands, Switzerland, Canada and Japan.^16^ Evaluation of the app is challenging as these decentralized apps do not collect information to protect privacy and therefore other sources need to be used. In Switzerland, 41 out of 6 380 (0.6%) confirmed cases reported the app (SwissCovid; launched June 2020) as the reason for the testing and a study estimated that 60% of app users who received a positive SARS-CoV-2 test triggered an app notification.^17,18^ Denmark’s app (Smitte|stop) has been downloaded by over 2 million times and with 53 000 infections reported since June 2020.^19^ Among the 87 904 people who identified the app as reason for testing, 814 tested positive (0.9%).^19^

It is essential to determine the optimal configurations of the app to identify relevant contacts as the time registered and the Bluetooth signal strength are affected by many factors. Several countries report their configurations settings and testing protocols. Testing in Switzerland showed that when attenuation is low (e.g. <50dBm), two devices were within a few meters, however higher attenuation values (50-70 dBm) offered less certainty and can result from devices that are up to 15 meters apart.^20^ Our study estimates the performance of the new Norwegian GAEN-based contact tracing app “Smittestopp v2” (open source) under relevant simulated real-life scenarios.^18^ We aimed at achieving a correct notification rate of ≥ 75% for close contacts.

## Methods

### Configurations for contact identification

Attenuation buckets divide the contact time registered by a phone into three group based on the Bluetooth attenuation registered by GAEN.^21^ The “low bucket” indicates the lowest attenuation of the Bluetooth signal (and thus closest contact), “high bucket” represents the highest attenuation and indicate larger distance. We based our initial configurations on internal testing and reports from other GAEN based apps (configuration 1); low bucket <57 dBm, middle bucket 57-63 dBm and high bucket >63 dBm. These settings were adjusted (configuration 2) after analyses of testing results; low bucket <57 dBm, middle bucket 57-68 dBm and high bucket >68 dBm. Each bucket was assigned a weight and the time spent in this bucket was multiplied by the weight. In addition, a threshold was set for this cumulative weight adjusted time in the buckets; all phones where the cumulative weighted time of all detected relevant exposures was above this time threshold would get an exposure notification.

### Devices used for testing

To test the GAEN configurations, we used 40 phones with a mix of android and iOS operating systems (50%) as well as a mix of brands and models (iPhone, Samsung, Nokia, Sony, Huawei, Xiaomi; see supplementary material). These phones represented the majority of phones used on the Norwegian market.^22^ A development version of the app was used to allow data collection on each phone, as well as the use of fake identifications to notify infection.

### Set-up of scenarios

We set up simulations of two scenarios where people are likely to be nearby unknown individuals in real life situations, namely standing in a queue and travelling with public transport. Each scenario included ten participants who carried two phones. They were asked to carry and use one phone in their hand and put one phone in their pocket. Of the ten participants, one person was identified as “infected” (tester 10), four people (testers 1, 2, 3 and 9) fell under the definition used for “close contact” in manual contact tracing (≤2 meters for ≥15 min) and five people did not fit this definitions (“other contacts”; either two-four meters away or <15 minutes within 2 meters) (Table 1).

**Table 1.** Overview of testers, their distance and duration to the “infected” individual; orange indicates “close contacts” and green indicated “other contacts”.

| Tester | Scenario 1 - queue |  | Scenario 2 - bus |  |
| --- | --- | --- | --- | --- |
| | $\leq 2$ meters | $> 2$ meters | $\leq 2$ meters | $> 2$ meters |
| 1 | 16 min. | - | 16 min. | - |
| 2 | 20 min. | - | 20 min. | - |
| 3 | 20 min. | - | 16 min. | - |
| 4 | 8 min. | 12 min. | 8 min. | - |
| 5 | - | 4 min. | - | 20 min. |
| 6 | - | 8 min. | 8 min. | - |
| 7* | - | 12 min. | 12 min. | - |
| 8 | - | 20 min. | 4 min. | - |
| 9 | 20 min. | - | 20 min. | - |

\* During one run of scenario 1, the “infected” phone was turned on too early resulting in a longer run and tester 7 was assigned as close contact.

### Scenario 1 – queue

In the queue scenario, one tester was situated in the position of a cashier and all other testers were positioned one meter apart in a queue (Figure 1A). After start of the scenario, everyone moved up one place closer to the cashier every four minutes and left the scenario after standing in front of the cashier. The total duration of this scenario was 20 min.

**Figure 1.**
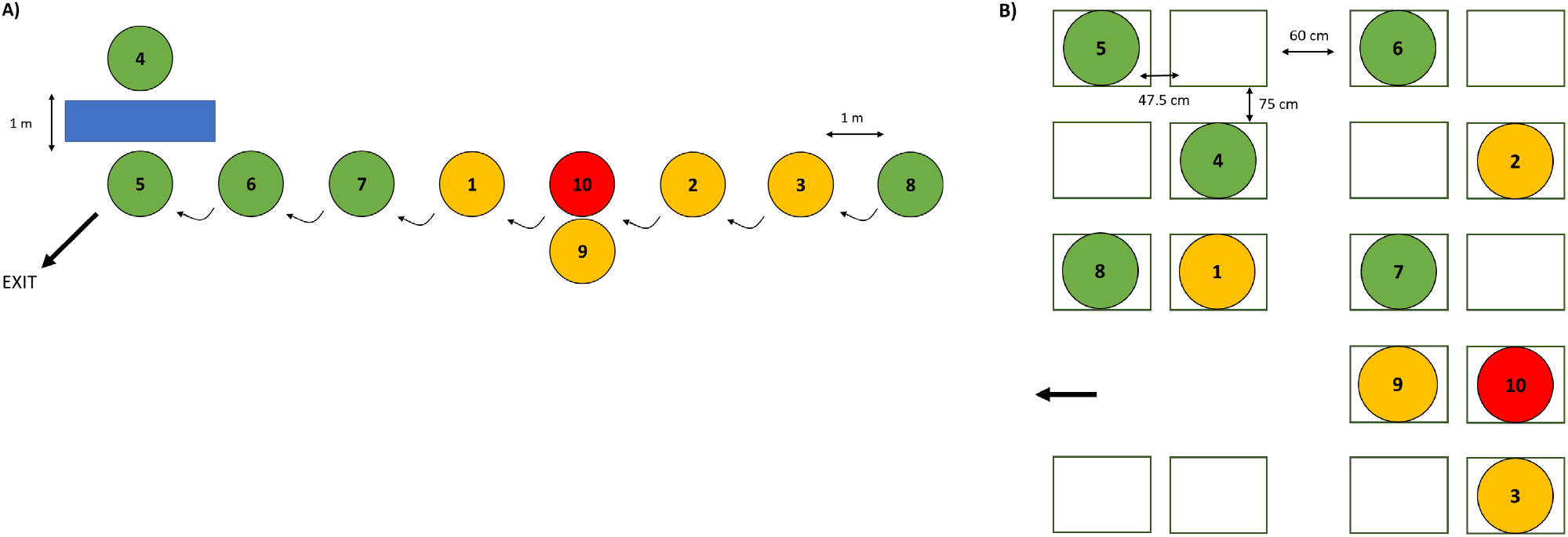
Schematic representation of the A) “queue scenario”, where participants simulated standing in a queue and B) “public transport” scenario, where participants simulated taking a bus to evaluate the notification rates via a contact tracing under various configuration settings. *Red: “infected” tester, “orange” tester within the definition of “close contact” and “green” tester not according to definition of “close contact”*.

### Scenario 2 – public transport

In the public transport scenario, we used the measurements of city buses as guidance, as provided by Ruter AS, the public transport authority for Oslo and Akershus counties in Norway. In this simulation, testers were sitting on chairs arranged according to the bus measurements and some were leaving the scenario every four minutes (Figure 1B). The total duration of the scenario was 20 minutes and the “infected” stayed in the bus during the whole scenario.

### Data collection and analyses

After notification of infection, data was pulled for each of the exposed phones (19 per run) and collected. The keys of the “infected” phone were used to check the data collected from the “exposed” phones. After data entry, the phones were cleaned, and the app reinstalled for the next test run.

We calculated the notification rates for all exposed phones included in the experiment using different weights for each of the buckets and time threshold to optimize the notification rates among “close contacts” and minimize the risk of incorrect notification of “other contacts”. We used a weight range between 1 and 2.5 for the low and middle buckets (high bucket is always 0) and time thresholds between 10 and 15 minutes (Table 3). In addition, we calculated differences in notification rates between phones in hand or in pocket, and android or iOS operating systems as well as the difference between other contact within 2 meters for <15 minutes compared to those further than 2 meters apart using χ^2^. All data analyses were performed in Stata/SE 16.0.

## Results

### Configuration 1

For configuration 1, we did four runs of scenario 1 and five runs of scenario 2, which included 171 exposed phones from nine infected phones. We had to exclude ten phones due to errors; three had the wrong date, one did not deactivated the app after ending the scenario, four phones ran on the incorrect operating system (iOS 13) and two phones were not activated in one of the runs.

Of the 161 exposed phones, 78 phones were of close contacts (48%) and 83 phones were of other contacts (52%). Overall, 69% (54) of close contacts and 45% (37) of other contacts registered time in the low or middle bucket and could therefore theoretically receive an exposure notification. The highest notification rate achieved was 63% among close contacts and 31% among other contacts, by using weights of 2.5 for the low and 2.0 for the middle bucket and a time threshold of 10 minutes (Table 2). With configuration 1, settings 4, 7 and 13 gave the best notification rates among close contacts (Table 2) and will be used for further analyses.

**Table 2.** Notifications among testers using different weight and time thresholds on configuration 1 of Smittestopp. Blue; settings with the highest notification rates among close contacts.

| Setting | Weights |  | Time threshold<br>minutes | Close contacts (78) |  | Other contacts (83) |  |
| --- | --- | --- | --- | --- | --- | --- | --- |
|  | Low bucket | Middle bucket |  | % | (n) | % | (n) |
| 1 | 2.5 | 1.0 | 10 | 60.3 | (47) | 20.5 | (17) |
| 2 |  |  | 13 | 55.1 | (43) | 15.7 | (13) |
| 3 |  |  | 15 | 51.3 | (40) | 13.3 | (11) |
| 4 | 2.5 | 1.5 | 10 | 61.5 | (48) | 22.9 | (19) |
| 5 |  |  | 13 | 60.3 | (47) | 19.3 | (16) |
| 6 |  |  | 15 | 55.1 | (43) | 16.9 | (14) |
| 7 | 2.5 | 2.0 | 10 | 62.8 | (49) | 31.3 | (26) |
| 8 |  |  | 13 | 61.5 | (48) | 20.5 | (17) |
| 9 |  |  | 15 | 60.3 | (47) | 19.3 | (16) |
| 10 | 2.0 | 1.0 | 10 | 60.3 | (47) | 19.3 | (16) |
| 11 |  |  | 13 | 51.3 | (40) | 14.5 | (12) |
| 12 |  |  | 15 | 48.7 | (38) | 12.1 | (10) |
| 13 | 2.0 | 1.5 | 10 | 61.5 | (48) | 22.9 | (19) |
| 14 |  |  | 13 | 56.4 | (44) | 19.3 | (16) |
| 15 |  |  | 15 | 53.9 | (42) | 13.3 | (11) |

Low bucket: <57dBm, Middle bucket: 57-68 dBm.
Other contacts are people that do not fit the definition of a close contacts; $\leq 2$ meters for >15 minutes.

**Table 3.**
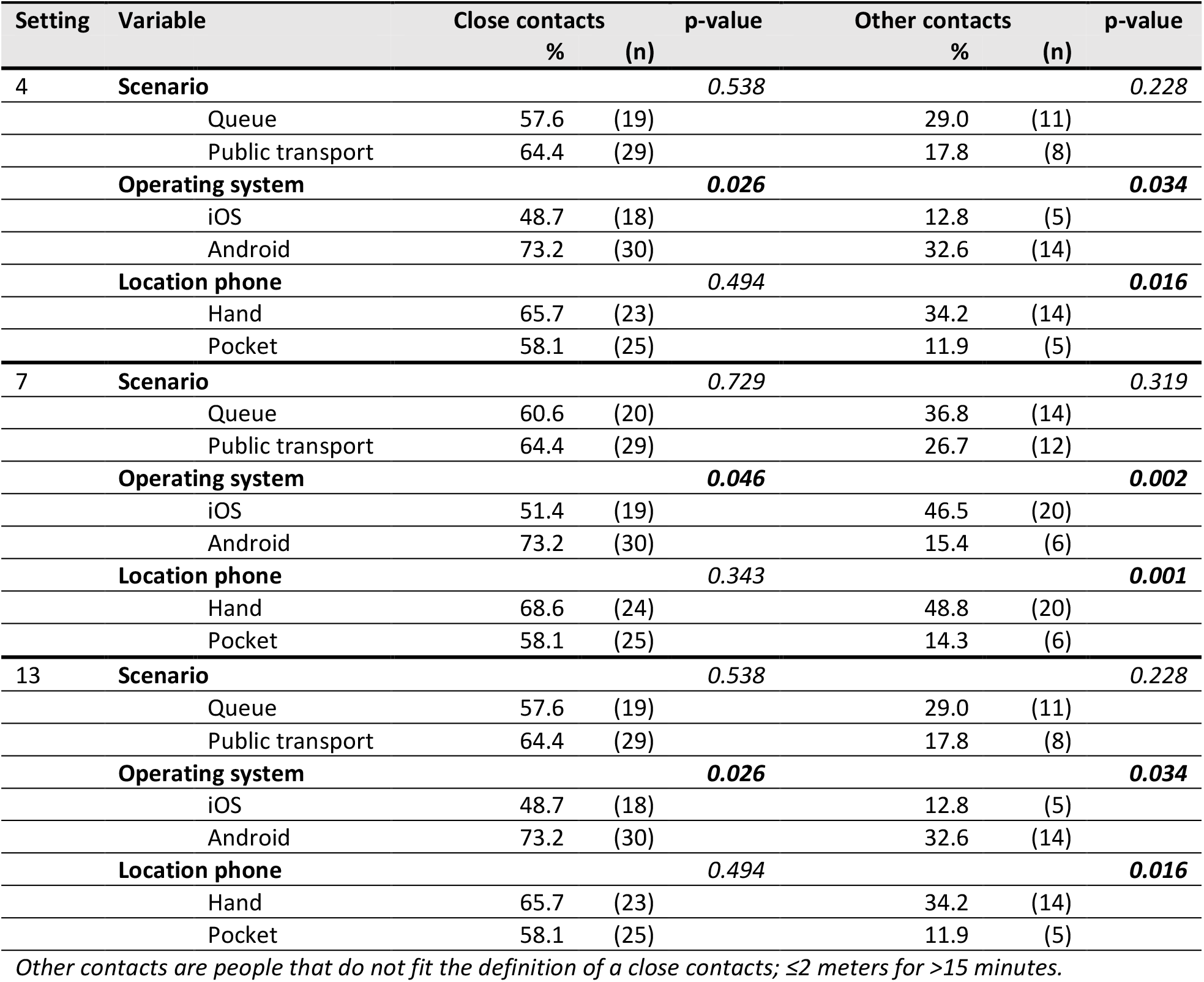
Notification rates among close contacts and other contacts using configuration 1 with selected settings for weights, split by various variables.

We found no significant differences between notification rates between the two scenarios (queue and public transport). Phones with iOS operating system had significantly lower notification rates among both close contacts and other contacts with all selected settings (Table 3). In addition, phones kept in the pocket had significantly lower notification rates among other contacts, but not among close contacts (Table 3).

As the highest notification among close contacts was lower than our acceptance criteria (75%), we adjusted the configuration of the buckets to have a higher upper limit for the middle bucket (from 63 to 68).

### Configuration 2

In total, we performed two runs of each scenario and included 76 exposed phones from four infected phones. We had to exclude three phones due to errors; one phone had an error submitting batches to the server and two phones ran on iOS 13.6.

Of the 73 exposed, 35 (48%) phones were of close contacts and 38 (52%) phones were of other contacts. Overall, 94.3% (33) of close contacts and 68.4% (26) of other contacts registered time in the low or middle bucket. Using configuration 2 resulted in significantly higher notification rates among close contacts than configuration 1 with all settings tested (p-value between 0.05 and 0.005). The most statistically significant difference was found with setting 7, where configuration 2 identified 88.6% of close contacts, compared to 62.8% with configuration 1 (p=0.005).

The highest notification rate among close contacts was 88.6% with setting 7, which also resulted in a high notification rate among other contacts (55.3%) (Table 4). Based on the best balance between high notification rate among close contacts and low notification rate among other contacts, we analyzed the data from setting 5, 6, 9 and 14 further (green in Table 4).

**Table 4.** Notifications among testers using different weight and time thresholds on configuration 2 of Smittestopp. Green; settings chosen to continue for further analyses.

| Setting | Weights |  | Time threshold<br>minutes | Close contacts (35) |  | Other contacts (38) |  |
| --- | --- | --- | --- | --- | --- | --- | --- |
|  | Low bucket | Middle bucket |  | % | (n) | % | (n) |
| 1 | 2.5 | 1.0 | 10 | 80.0 | (28) | 39.5 | (15) |
| 2 |  |  | 13 | 74.3 | (26) | 26.3 | (10) |
| 3 |  |  | 15 | 71.4 | (25) | 23.7 | (9) |
| 4 | 2.5 | 1.5 | 10 | 85.7 | (30) | 44.7 | (17) |
| 5 |  |  | 13 | 80.0 | (28) | 34.2 | (13) |
| 6 |  |  | 15 | 80.0 | (28) | 34.2 | (13) |
| 7 | 2.5 | 2.0 | 10 | 88.6 | (31) | 55.3 | (21) |
| 8 |  |  | 13 | 85.7 | (30) | 39.5 | (15) |
| 9 |  |  | 15 | 82.9 | (29) | 39.5 | (15) |
| 10 | 2.0 | 1.0 | 10 | 80.0 | (28) | 39.5 | (15) |
| 11 |  |  | 13 | 71.4 | (25) | 26.3 | (10) |
| 12 |  |  | 15 | 71.4 | (25) | 23.7 | (25) |
| 13 | 2.0 | 1.5 | 10 | 85.7 | (30) | 44.7 | (17) |
| 14 |  |  | 13 | 80.0 | (28) | 34.2 | (13) |
| 15 |  |  | 15 | 77.1 | (27) | 34.2 | (13) |

*Low bucket: <57dB, Middle bucket: 57-68 dB*
*Other contacts are people that do not fit the definition of a close contacts; $\leq 2$ meters for $>15$ minutes.*

We found no significant differences between notification rates in the two scenarios (queue and public transport). Phones with an iOS operating system or phones that were placed in pockets had lower notification rates with all selected settings (Table 5).

**Table 5.** Notification rates among close contacts and other contacts using configuration 2 with selected settings for weights, split by various variables.

| Setting | Variable | Close contacts |  |  | Other contacts |  |  |
| --- | --- | --- | --- | --- | --- | --- | --- |
|  |  | % | (n) | p-value | % | (n) | p-value |
| 5 / 6 | Scenario |  |  | 0.612 |  |  | 0.732 |
|  | Queue | 76.5 | (13) |  | 31.6 | (6) |  |
|  | Public transport | 83.3 | (15) |  | 36.8 | (7) |  |
|  | Operating system |  |  | 0.062 |  |  | 0.087 |
|  | iOS | 68.4 | (13) |  | 21.1 | (4) |  |
|  | Android | 93.8 | (15) |  | 47.4 | (9) |  |
|  | Location phone |  |  | 0.042 |  |  | 0.207 |
|  | Hand | 94.1 | (16) |  | 44.4 | (8) |  |
|  | Pocket | 66.7 | (12) |  | 25 | (5) |  |
| 9 | Scenario |  |  | 0.330 |  |  | 0.740 |
|  | Queue | 76.5 | (13) |  | 42.1 | (8) |  |
|  | Public transport | 88.9 | (16) |  | 36.8 | (7) |  |
|  | Operating system |  |  | 0.117 |  |  | 0.020 |
|  | iOS | 73.7 | (14) |  | 21.1 | (4) |  |
|  | Android | 93.8 | (15) |  | 57.9 | (11) |  |
|  | Location phone |  |  | 0.009 |  |  | 0.054 |
|  | Hand | 100 | (17) |  | 55.6 | (10) |  |
|  | Pocket | 66.7 | (12) |  | 25 | (5) |  |
| 14 | Scenario |  |  | 0.612 |  |  | 0.732 |
|  | Queue | 76.5 (13) |  |  | 31.6 | (6) |  |
|  | Public transport | 83.3 (15) |  |  | 36.8 | (7) |  |
|  | Operating system |  |  | 0.062 |  |  | 0.087 |
|  | iOS | 68.4 (13) |  |  | 21.1 | (4) |  |
|  | Android | 93.8 (15) |  |  | 47.4 | (9) |  |
|  | Location phone |  |  | 0.042 |  |  | 0.207 |
|  | Hand | 94.1 (16) |  |  | 44.4 | (8) |  |
|  | Pocket | 66.7 (12) |  |  | 25.0 | (5) |  |

*Other contacts are people that do not fit the definition of a close contacts; $\leq 2$ meters for $>15$ minutes.*

Among other contacts, people who were within 2 meters for less than 15 minutes had a significantly higher notification rate than people further (2-4 meters) away (Table 6).

**Table 6.** Notification rates among individuals who do not fit the definition of a close contact (≤2 meters for >15 minutes.) divided by distance from “infected” phone using configuration 2.

| Setting | $\leq 2$ meters | | $> 2$ meters | | p-value |
| --- | --- | --- | --- | --- | --- |
|  | % | (n) | % | (n) |  |
| 5 / 6 | 66.7 | (8) | 19.2 | (5) | 0.004 |
| 9 | 75.0 | (9) | 23.1 | (6) | 0.002 |
| 14 | 66.7 | (8) | 19.2 | (5) | 0.004 |

## Discussion

In this study we optimized the precision of the GAEN-based Norwegian contact tracing app Smittestopp under “real-life” scenarios to target close contacts. Our results show that performance could be considerably improved by adjusting the configurations of the buckets. As expected, we observed variations between different devices with phones with the iOS operating system generally having a lower sensitivity than those with Android as well as those in the pocket. This supports previous knowledge on the inaccuracy of using Bluetooth-based signal in proximity estimation and is an important aspect to take into consideration when communicating advice to the public.^11,23^

The success of Smittestopp as a tool to control national outbreaks relies on its ability to timely and correctly identify and notify those exposed as well as having a high adoption rate in the population. Although difficult to model, others have suggested an adoption rate of above 20% already has an impact on infection rates.^24^ The decision to recommend testing as soon as possible was deemed less of a burden than recommending quarantine and therefore expected to increase uptake and acceptance of the application. Since local storage of data do not allow, identification and direct follow up of cases and contacts via Smittestopp can only function as a supplement to, and not replace, manual contact tracing conducted by the local health authorities. Thus, to maximize the benefit of this tool, it is important to reach those who might get exposed in public spaces where manual contact tracing is particularly challenging.^25^ Furthermore, it is important to reach subgroups of the public (age groups, nationalities) being overrepresented among covid-19 cases and living in areas where the incidence rate has remained high throughout the pandemic.^26^ In Norway, over half of the phones run on iOS and therefore this can affect the notification rates. However, the settings chosen were based on the average notifications rates, which include 50% phones running on iOS to represent the Norwegian setting.

Many countries have implemented a contact tracing app based on the GAEN solution, but very few have published data on configuration testing in peer reviewed journals. Germany has shown the scenarios they used for testing on their github, notification rates were among close contacts was 47% and 8% among “non close” contacts with bucket thresholds of 55 and 63 dBm.^27^ These results are very similar to those in this study using configuration 1 settings. The Netherlands reported on a field testing with seven scenarios to decide the optimal settings to identify exposure within 1.5 meters for at least 15 minutes. These tests identified that the cut-off value should be between 68 and 75dBm and the time threshold should be less than 15 minutes for closer contacts. Therefore, they included all contacts with an attenuation ≤63 dBm for >10 minutes, or 64-73dBm for >15 minutes.^28^ Considering that the Netherlands uses risk scores instead of buckets, meaning they will identify exposure based on single contacts, these results are in line with the settings chosen in Norway. Based on their scenario testing, Switzerland has a configuration which is stricter than in Norway; low bucket ≤55dBm weighted 1.0, medium bucket 55-63 dBm with a weight of 0.5 and a time threshold of 15 minute over a calendar day. They showed that 88% of phones within 2 meters had an attenuation below 63 dBm, but it is unclear how the duration is factored in. An upper threshold of 68 dBm, as chosen by Norway, would result in 98% of those within 2 meters recording an attenuation of 68 dBm compared with 94% in this study. However, the data presented does not show how it is related to duration and identification of relevant contacts. Our study adds to the current literature by showing the influence of changing the configuration settings on notification of relevant close contacts and other contacts, the considerations taken into account when choosing the settings, as well as some of the factors affecting exposure identification such as operating system of the phone and the location of the phone during exposure by using real-life scenarios with different phones ^20^.

In Norway, contact tracing is normally initiated to identify and quarantine those who have been within 2 meters for over 15 minutes to a confirmed COVID-19 case during the infectious phase of the disease.^2^ However, the true risk of being infected depends on a combination of factors related to the host, the exposed, the exposure situation and environment as well as the virus itself. During manual contact tracing, health care professionals evaluate the risk and will recommend quarantine and/or testing based on this assessment. With digital apps, such as Smittestopp, notifications need to be sent based on a set of criteria defined by the app, namely duration of contact, Bluetooth attenuation, and when exposure took place. Due to Bluetooth attenuation as well as the duration being affected by many factors, these digital contact tracing apps are not specific in sending only notifications to close contacts.^11,20,27,28^ The decisions on the configuration settings will therefore always be a balance between sending notifications to “true” close contacts and limiting notifications sent to other contacts as well as potential recommendations, which can be affected by testing capacity. The decision on the current configuration settings was based on a combination of variables; 1) Notification rate among close contacts, 2) notification rate among other contacts and 3) the type of advice and measures for contacts. We adjusted the thresholds of the buckets to achieve a correct notification rate of minimum 75%. In addition, closer proximity was deemed a higher risk factor than duration and therefore we accepted higher notification rates for those within 2 meters for less than 15 minutes than among those further away. Recommending quarantine for contacts identified via the app was considered to not be a proportionate measure, based on the sensitivity of Bluetooth technology as well as unknown last day of exposure in the current solution. In Norway, the current advice is to get tested as soon as possible and stay home until the test result is ready. Based on current testing capacity, people will be able to get tested and receive their results within 24-48 hours in most of the country, which would not be a large personal burden and therefore a higher notification rate among other contacts was acceptable. In addition, a study from Switzerland showed that app notified contacts went into quarantine earlier than those without app notification ^10^, identifying that digital tools can also contribute to a quicker response among known contact.

In conclusion, we show that the accuracy of the app could be considerably improved by adjusting the GAEN-configurations. These findings provide guidance to health authorities on the expected notification rates and limitations of app-based contact tracing. Experimental data on the performance of the app under real-life conditions could help building confidence among the public as well as push the technological process and improvement forward. These configurations are easily adjustable and should be regularly reassessed based on a combination of factors which could change over time, such as disease prevalence, infectiousness of new virus variants as well as changes in national advice and control measures. Thus, configurations settings should be carefully adapted to the national situation and tested under relevant exposure scenarios and not copied from other existing solutions abroad. Although there are still technological and other limitations that needs to be overcome before GAEN-based apps could replace manual contact tracing, we believe that transparency around the development and testing could contribute to increased acceptability and trust among the public.

## Data Availability

After publication in a peer reviewed journal, the data will be made available online.

## Supplementary material

Overview of phones used in scenario testing of Smittestopp v2

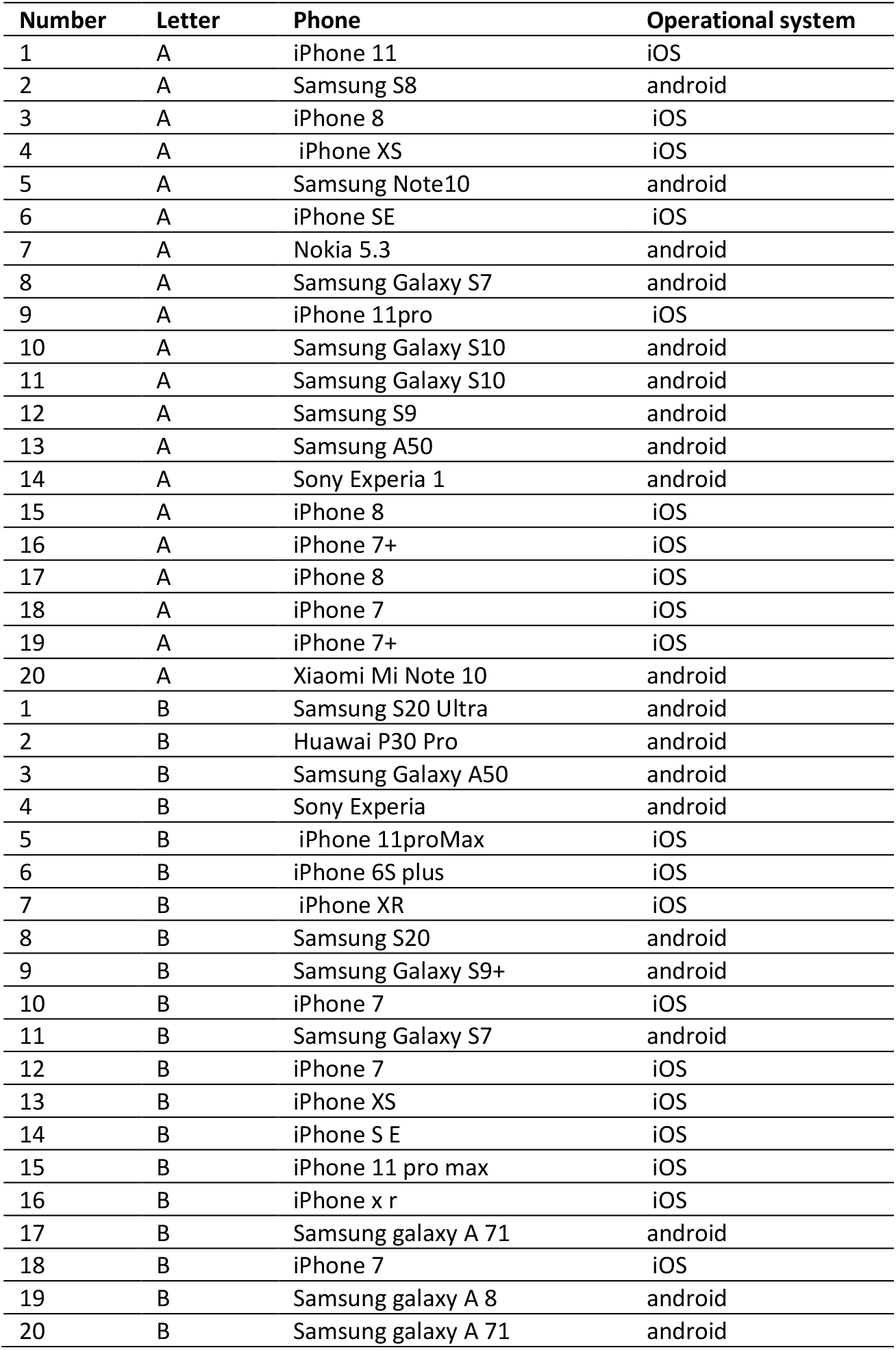

| Number | Letter | Phone | Operational system |
| --- | --- | --- | --- |
| 1 | A | iPhone 11 | iOS |
| 2 | A | Samsung S8 | android |
| 3 | A | iPhone 8 | iOS |
| 4 | A | iPhone XS | iOS |
| 5 | A | Samsung Note10 | android |
| 6 | A | iPhone SE | iOS |
| 7 | A | Nokia 5.3 | android |
| 8 | A | Samsung Galaxy S7 | android |
| 9 | A | iPhone 11pro | iOS |
| 10 | A | Samsung Galaxy S10 | android |
| 11 | A | Samsung Galaxy S10 | android |
| 12 | A | Samsung S9 | android |
| 13 | A | Samsung A50 | android |
| 14 | A | Sony Experia 1 | android |
| 15 | A | iPhone 8 | iOS |
| 16 | A | iPhone 7+ | iOS |
| 17 | A | iPhone 8 | iOS |
| 18 | A | iPhone 7 | iOS |
| 19 | A | iPhone 7+ | iOS |
| 20 | A | Xiaomi Mi Note 10 | android |
| 1 | B | Samsung S20 Ultra | android |
| 2 | B | Huawei P30 Pro | android |
| 3 | B | Samsung Galaxy A50 | android |
| 4 | B | Sony Experia | android |
| 5 | B | iPhone 11proMax | iOS |
| 6 | B | iPhone 6S plus | iOS |
| 7 | B | iPhone XR | iOS |
| 8 | B | Samsung S20 | android |
| 9 | B | Samsung Galaxy S9+ | android |
| 10 | B | iPhone 7 | iOS |
| 11 | B | Samsung Galaxy S7 | android |
| 12 | B | iPhone 7 | iOS |
| 13 | B | iPhone XS | iOS |
| 14 | B | iPhone S E | iOS |
| 15 | B | iPhone 11 pro max | iOS |
| 16 | B | iPhone x r | iOS |
| 17 | B | Samsung galaxy A 71 | android |
| 18 | B | iPhone 7 | iOS |
| 19 | B | Samsung galaxy A 8 | android |
| 20 | B | Samsung galaxy A 71 | android |

